# Cerebello-thalamic activity drives an abnormal motor network into dystonic tremor

**DOI:** 10.1101/2021.11.03.21265803

**Authors:** Freek Nieuwhof, Ivan Toni, Michiel F. Dirkx, Cecile Gallea, Marie Vidailhet, Arthur W.G. Buijink, Anne-Fleur van Rootselaar, Bart P.C. van de Warrenburg, Rick C. Helmich

## Abstract

Dystonic tremor syndromes are highly burdensome and treatment is often inadequate. This is partly due to poor understanding of the underlying pathophysiology. Several lines of research suggest involvement of the cerebello-thalamo-cortical circuit and the basal ganglia in dystonic tremor syndromes, but their role is unclear. Here we aimed to investigate the contribution of the cerebello-thalamo-cortical circuit and the basal ganglia to the pathophysiology of dystonic tremor syndrome, by directly linking tremor fluctuations to cerebral activity during scanning. In 27 patients with dystonic tremor syndrome (dystonic tremor: *n*=23; tremor associated with dystonia: *n*=4), we used concurrent accelerometery and functional MRI during a posture holding task that evoked tremor, alternated with rest. Using multiple regression analyses, we separated tremor-related activity from brain activity related to (voluntary) posture holding. Using dynamic causal modelling, we tested for altered effective connectivity between tremor-related brain regions as a function of tremor amplitude fluctuations. Finally, we compared grey matter volume between patients (*n*=27) and matched controls (*n*=27).

We found tremor-related activity in sensorimotor regions of the bilateral cerebellum, contralateral ventral intermediate (VIM) and ventro-oralis posterior nuclei (VOp) of the thalamus, contralateral primary motor cortex (hand area), contralateral pallidum, and the bilateral frontal cortex (laterality with respect to the tremor). Grey matter volume was increased in patients compared to controls in the portion of contralateral thalamus also showing tremor-related activity, as well as in bilateral medial and left lateral primary motor cortex, where no tremor-related activity was present. Effective connectivity analyses showed that inter-regional coupling in the cerebello-thalamic pathway, as well as the thalamic self-connection, were strengthened as a function of increasing tremor power.

These findings indicate that the pathophysiology of dystonic tremor syndromes involves functional and structural changes in the cerebello-thalamo-cortical circuit and pallidum. Deficient input from the cerebellum towards the thalamo-cortical circuit, together with hypertrophy of the thalamus, may play a key role in the generation of dystonic tremor syndrome.

## 1. Introduction

Postural tremor is a highly burdensome symptom that is present in 17-55% of patients with dystonia^1-3^. Tremor is defined as an involuntary, rhythmic, oscillatory movement of a body part^4^. Two types of dystonic tremor syndrome are distinguished: dystonic tremor, defined as tremor in a body part affected by dystonia, and tremor associated with dystonia, defined as tremor in a non-dystonic body part. Clinically, dystonic tremor syndrome typically involves an action tremor (but it may also occur during rest), tremor occurrence can be position or task-specific, and the tremor often has an irregular aspect^5^. Current treatments, such as deep brain stimulation (DBS), pharmacological interventions, or botulinum toxin, often have unsatisfactory results^6,7^. Precise knowledge about the underlying pathophysiology of dystonic tremor syndromes is lacking, preventing mechanism-based treatments. However, to date, only few studies have investigated the pathophysiology of dystonic tremor syndromes, and no studies focused on tremor-related brain activity.

Dystonic tremor syndromes may partly share pathophysiological mechanisms with dystonia itself, or with tremor syndromes that are phenotypically similar, such as essential tremor^8^. One possibility is that the pathophysiology of dystonic tremor syndrome involves the cerebello-thalamo-cortical circuit and its connections to the basal ganglia^8,9^. This hypothesis is supported by several lines of research. Stereotactic interventions on the ventral intermediate nucleus of the thalamus (VIM), a cerebellar relay nucleus, as well as DBS of the internal globus pallidus (GPi) or its thalamic relay nucleus (the ventro-oralis posterior (VOp), can both suppress tremor in dystonia^7,10-14^. Furthermore, functional MRI during a grip-force task has been used as a proxy of tremor-related cerebral activity^15^, showing similar grip-force related activity in the cerebellum between dystonic tremor and essential tremor. Since cerebellar dysfunction is well-established in essential tremor^16-19^, this suggests that cerebellar dysfunction may also play a role in dystonic tremor syndromes. In the same study, dystonic tremor was associated with reduced functional connectivity of the VIM, GPi, and dentate nucleus^15^. In two other functional MRI studies, patients with dystonic voice tremor had increased cerebellar activity during speech production^20^ and reduced self-inhibition of the putamen during rest^21^. However, these previous studies did not directly relate altered brain activity to tremor, which limits the conclusions that can be drawn. In a structural MRI study, dystonic tremor patients had an increased grey matter volume (GMV) in the sensorimotor cortex when compared to controls and essential tremor patients^22^, hinting at cortical rather than subcortical mechanisms. Finally, classical eye blink conditioning and temporal somatosensory discrimination, which are markers for cerebellar and basal ganglia dysfunction, respectively, are affected in patients with dystonic tremor syndromes^23,24^. Taken together, these findings suggest that both the cerebello-thalamo-cortical circuit and the basal ganglia are involved in dystonic tremor syndromes, but direct evidence is lacking, and the role of each circuit remains unclear.

Here, we aimed to identify the cerebral circuit underlying dystonic tremor syndrome, by focusing on specific patterns of tremor-related activity independent of voluntary movements (such as posture holding). We combined accelerometery with functional MRI, using the same method as previously applied to Parkinson’s disease tremor^25,26^, Holmes tremor^27^ and essential tremor^16,17^. We disentangled cerebral activity related to hand lifting, posture holding, and hand lowering from cerebral activity related to fluctuations in tremor amplitude. We hypothesized to find tremor-related activity in the cerebello-thalamo-cortical circuit and the basal ganglia (specifically, the GPi), possibly as a function of structural changes and/or clinical tremor characteristics. Finally, within the tremor-related network, we investigated the role of inter-regional connections in the generation of dystonic tremor syndrome. To this end, we used dynamic causal modelling to test where cerebral activity first arises during hand-lifting, and which connections are related to fluctuations in tremor amplitude.

## 2. Materials and methods

### 2.1. Participants

We included 27 patients with dystonic tremor syndrome from the Radboud university medical centre (Radboudumc Nijmegen, 18 patients), Academic Medical Center (AMC Amsterdam, 7 patients), Maastricht University Medical Center+ (MUMC Maastricht, 1 patient) and Canisius Wilhelmina Ziekenhuis (CWZ Nijmegen, 1 patient). For comparison of grey matter volume with patients, we selected 27 healthy controls from two prior studies that best matched our dystonic tremor syndrome group on age and sex^28,29^. The healthy controls were 61.0 ± 11.5 years old (range 34-80) and sex was distributed as in dystonic tremor syndrome patients (13 male, 14 female). Inclusion criteria for dystonic tremor syndrome patients were a clinical diagnosis of dystonic tremor or tremor associated with dystonia, with the presence of primary focal or segmental dystonia, according to the most recent consensus statement^4^. Patients with questionable dystonia and bilateral, sinusoidal, highly regular (in amplitude and rhythm) postural tremor were classified as essential tremor plus and excluded from this study^4,30^.

Exclusion criteria for all participants were: MRI contraindications, a history of traumatic brain injury or stroke, moderate to severe head tremor when lying supine, cognitive dysfunction (defined as a clinical diagnosis of mild cognitive impairment or dementia), and the use of anti-tremor medication other than propranolol. All participants participated voluntarily and gave their written informed consent prior to starting the study. The study was approved by the ethical committee ‘Commissie mensgebonden onderzoek (CMO) regio Arnhem-Nijmegen’ and was performed according to the principles of the Declaration of Helsinki. Propranolol was tapered off for the patients using it (*n*=10) in the week prior to measurements so all measurements were done in the off-medication state. To prevent any effect of botulinum toxin treatment, measurements for the two patients who received injections in the most tremulous arm were scheduled three and six months after the latest injection.

### 2.2. Experimental procedures and paradigm

Patients visited our lab on two separate sessions. During the first visit, we performed a clinical examination and videotaped this for later review and confirmation of diagnosis by two experienced neurologists (RH and BvdW). Table 1 provides details on the clinical testing. Using functional MRI, we scanned participants during both the first and the second visit (*n*=15) or during one of the visits (*n*=12). During scanning, we used a block design to evoke postural tremor while allowing sufficient periods of rest (20 blocks of 30 seconds posture holding alternated with 20 blocks of 9-11 seconds rest, in total 13.3 minutes). Specifically, we asked participants to rest their hands and arms on their hip or the scanner bed whenever the text ‘RUST’ (Dutch for rest) was displayed. When ‘STREK’ (Dutch for stretch) was displayed, we asked participants to assume an individually defined tremor-evoking posture with the arm that was most affected by tremor.

**Table 1.** Characteristics of participants.

|  | Dystonic tremor syndrome (N=27) |
| --- | --- |
| | mean ± std (range) |
| General |  |
| Age (years) | 62.0 ± 12.7 (28.0 - 82.0) |
| Sex (M/F) | 13/14 |
| FAB | 16.2 ± 1.9 (11.0 - 18.0) |
| MOCA | 26.8 ± 2.9 (19.0 - 31.0) |
| Tremor |  |
| Most affected arm (left/right) | 11/16 |
| Postural tremor frequency (Hz) | 5.7 ± 1.6 (3.8 - 9.4) |
| TRS total score | 41.8 ± 15.8 (18.0 - 84.0) |
| TRS most affected hand | 17.0 ± 5.3 (8.0 - 26.0) |
| Tremor duration (years) | 24.1 ± 15.6 (8.0 - 58.0) |
| Tremor response to alcohol (0=no response, 100=full tremor suppression) | 19.6 ± 28.1 (0.0 - 100.0) |
| Propranolol usage for tremor (yes/no) | 10/17 |
| Head tremor (yes/no) | 9/18 |
| Rest tremor (yes/no) | 25/2 |
| Kinetic tremor (yes/no) | 27/0 |
| Irregular tremor (yes/no) | 16/11 |
| Asymmetric tremor (yes/no) | 20/7 |
| Position-dependent tremor (yes/no) | 18/9 |
| Unilateral tremor (yes/no) | 3/24 |
| Dystonia |  |
| BFM total score | 7.9 ± 6.7 (0.5 - 27.5) |
| Cervical dystonia (yes/no) | 23/4 |
| Most tremulous arm dystonia (yes/no) | 23/4 |
| Least tremulous arm dystonia (yes/no) | 17/10 |
| Speech/swallow dystonia (yes/no) | 5/22 |
| Eyes dystonia (yes/no) | 1/26 |
| Leg dystonia (yes/no) | 2/25 |
| Trunk dystonia (yes/no) | 7/20 |
| Mouth dystonia (yes/no) | 0/27 |

The Fahn-Tolosa-Marin Tremor Rating Scale (TRS) parts A, B and C were assessed for clinical tremor severity (possible ranges 0-88, 0-36 and 0-28<sup>52</sup>. The sum score for items covering the most tremulous hand was calculated and used as measure for tremor severity of the most affected hand (possible range 0-28). Peak tremor frequency and the effect of weighing were assessed by using accelerometry. The Burke-Fahn-Marsden rating scale (BFM, possible range 0-120) was used for severity of dystonia<sup>53</sup>, the Montreal Cognitive Assessment (MOCA, possible range 0-31) score for general cognitive functioning<sup>54,55</sup>, the Frontal Assessment Battery (FAB, possible range 0-18) for frontal lobe functioning<sup>56,57</sup>.

### 2.3. Tremor analysis

During scanning, we recorded tremor by using an MRI-compatible tri-axial accelerometer (Brain Products; sampling frequency *(Fs)* = 5kHz)^31^. We placed the accelerometer on the location where tremor was best captured (dorsum of the hand or one of the fingers). Accelerometery data was detrended and demeaned. We then segmented the data into 5s segments over which we calculated power spectra with 0.2 Hz spectral resolution. These power spectra were averaged and used to determine the channel and frequency with highest tremor power. We then calculated the time-frequency representation of this channel between 2 and 18 Hz. For this, we used a Hanning taper with a window length equal to 8 periods at peak tremor frequency (e.g. 2s for 4Hz tremor) to minimize spectral leakage. From this, we extracted the tremor power time-course at each individual’s peak tremor frequency (Fig 1). This time course was down sampled to the repetition time to obtain scan-to-scan tremor power.

**Figure 1.**
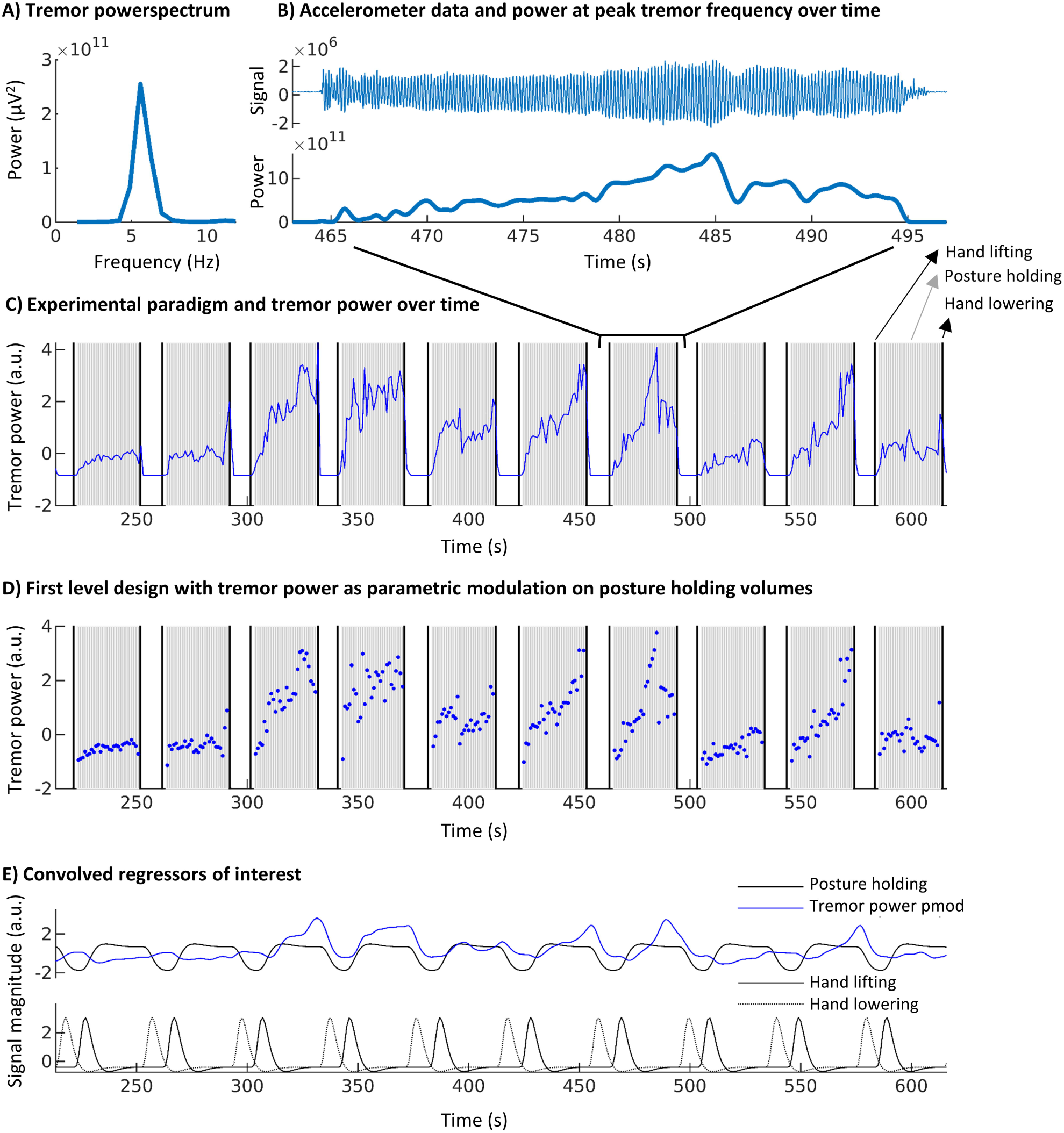
Tremor analysis and first level design. This figure illustrates the implementation of postural tremor analyses in our fMRI design for a representative participant. **(A)** Average power spectrum over the entire task, used to determine peak tremor **frequency (B)** Accelerometery signal of the axis with highest peak tremor power (top) and corresponding power over time at peak tremor frequency (bottom) for a single block. **(C)** Tremor power over time (blue) plotted over the experimental paradigm that included hand lifting, posture holding and hand lowering (10 out of 20 blocks). Note that, since tremor was present during posture holding and absent during rest, tremor power over time and posture holding are positively correlated. **(D)** Implemented first level design which mathematically separated posture holding and tremor power over time. Posture holding was modelled per volume with tremor power as parametric modulation per volume. **(E)** Regressors of interest in the first level design after convolution with the hemodynamic response function. Note that posture holding and tremor power are no longer positively correlated. This model allows for determination of tremor-related activity independent from hand onset, posture holding and hand offset movements. Pmod = parametric modulation. a.u. = arbitrary units.

### 2.4. MRI image acquisition and pre-processing

MRI images were acquired on a Siemens PRISMA 3T MRI system, using a 64-channel head-neck coil. T2*-weighted images were obtained using multiband echo planar imaging (EPI), with multiband acceleration factor 6, repetition time 1s, echo time 34 ms, 2.0 mm isometric voxels, 72 slices and a field of view of 210 mm. A high-resolution anatomical image was acquired using an MP-RAGE (magnetization-prepared rapid gradient-echo) sequence for both patients and healthy controls (repetition time 2300 ms, echo time 3.03 ms, voxel size 1.0 mm isometric, 192 sagittal slices, field of view 256 mm). Functional images were pre-processed using previously described procedures with a net smoothing kernel of 6 mm^26^. This included ICA-AROMA (independent component analysis-based automatic removal of motion artefacts) for automatic classification and removal of noise components^32,33^. Since ICA-AROMA was developed for resting-state data, the automatic classification was manually checked and corrected to prevent signal at task or tremor frequency to be inadequately classified as noise. We added movement parameters to our first level design to ensure this procedure did not reintroduce movement artefacts, which can resemble signal of interest. We normalized functional images to MNI (Montreal Neurological Institute) space and, for optimal sensitivity in the cerebellum, to a cerebellum specific template using the SUIT toolbox^34^.

### 2.5. Functional MRI analysis

For each participant, we performed a multiple regression analysis at the first level using the general linear model implemented in SPM12 (Fig 1). For optimal timing, we manually defined the time points of hand lifting and lowering based on the raw accelerometery data. We used these time points to model hand lifting and lowering as stick functions. In addition, we modelled posture holding (maintaining the tremor-evoking arm position) per volume, starting 2s after arm lifting and ending 1s before arm-lowering. Specifically, we entered onsets at times of volume initiation and durations equal to the repetition time (1s). We modelled fluctuations in tremor power by adding scan-to-scan tremor power as parametric modulation to these posture holding volumes. This allowed us to detect fluctuations in cerebral activity associated with fluctuations in tremor power, independent from cerebral activity related to posture holding, hand lifting or hand lowering. A similar approach has been used before^26^. For removal of non-neural noise and motion artefacts we added the average time course of the bilateral ventricles and 36 movement parameters (Volterra expansion: translation and rotation of 3 axes, original and first derivative, linear, quadratic and cubic polynomials) as nuisance regressors^35^. We averaged over sessions at the first level for participants who performed the task during both sessions (N=15, average time between sessions 5.5 ± 3.8 months). Two sessions with a maximum scan-to-scan displacement >3mm were excluded from analyses (for two participants who both had another session with less displacement). Mean scan-to-scan displacement was 0.14 ± 0.07 mm (range 0.06-0.37mm) and maximal scan-to-scan displacement was 0.94 ± 0.68 mm (range 0.21-2.64mm). Parameter estimates for all regressors were obtained by restricted maximum-likelihood estimation, and a temporal high pass filter with 128 s cut-off was used.

First-level contrast images were taken to the second level and entered into one-sampled t-tests. For participants with most severe tremor at the left hand (N=11), we flipped the contrast images in the axial plane (both for whole brain and cerebellum specific (SUIT) analyses)^25,31^. In this way, the primary motor cortex contralateral to the tremulous arm was always on the left side in the image.

### 2.6. Effective connectivity: Dynamic causal modelling

The analyses above demonstrated tremor-related activity in nodes of the cerebello-thalamo-cortical circuit (cerebellum, thalamus and BA4) and basal ganglia (GPi). However, it does not specify how these nodes interact with each other (effective connectivity), and most importantly, how fluctuations in inter-regional coupling are related to fluctuations in tremor amplitude. To investigate this, we used Dynamic Causal Modelling (DCM), which is a Bayesian method of inference where one defines one or more cerebral models based on predefined hypotheses to test for causal influences that one neural system exerts over the other^36^. Here, we constructed a model space to test: (1) where cerebral activity first arises during hand lifting (aim 1), and (2) which (self)-connections are related to fluctuations in tremor amplitude (aim 2). This approach has been used before for Parkinson’s disease tremor (aim 1)^31^ and for essential tremor (aim 2)^17^. We used a model with four regions of interests with tremor-related activity: sensorimotor areas of the cerebellum^37^, the thalamus, motor cortex (BA4), and GPi (see statistical analysis paragraph for exact ROI definition). We extracted BOLD fMRI timeseries using the first eigenvariate of voxels showing tremor related-activity in these ROIs, adjusting for other effects (i.e. hand lifting, posture holding, tremor, and hand lowering). All intrinsic connections (DCM.A) were based on direct anatomical connections as described in previous work^26,31,38^. Given that tremor-related activity in the thalamus overlapped both VIM and VOp, and given that our spatial resolution was insufficient to reliably disentangle these two thalamic nuclei, we considered both cerebellum-thalamus (VIM) and GPi-thalamus (VOp) connections in our model (Fig 3b). Next, we added hand lifting as a direct input (DCM.C) on the four nodes and tremor amplitude fluctuations (parametric modulator) as modulatory input (DCM.B) on the intrinsic interregional and self-connections. In DCM, self-connections reflect self-inhibition, which is incorporated to ensure decay of activity in the absence of input^36^. This yielded a total of 4 (direct input on 4 possible nodes) x 2^11^ (modulatory input on intrinsic connections) = 8,192 models, of which we defined four model families that shared a unique direct input (DMC.C) to one of the four nodes^39^. We used deterministic DCM 12.5 for all our analyses.

We used random effects Bayesian model selection to determine which of the four model families most likely generated the observed BOLD responses^40^. Next, we applied Bayesian model averaging to calculate mean parameters of the winning model family considering the relative model evidence^39,40^. We tested DCM.B parameters using one-sample two-tailed t-tests against zero (FDR corrected for 11 comparisons) to investigate significant associations between effective connectivity and tremor amplitude fluctuations. Significant parameters were correlated with clinical tremor severity (sum score of TRS items of the most affected hand) and dystonia severity (Burke-Fahn-Marsden; BFM) using Spearman’s Rho.

### 2.7. Structural MRI analysis

We tested for structural brain changes in dystonic tremor syndrome patients using voxel-based morphometry (VBM). Structural images were segmented into grey matter, white matter, and cerebrospinal fluid. Grey matter images were normalized to Montreal Neurological Institute (MNI) space and the Cerebellar template (SUIT) via a study specific template by using Diffeomorphic anatomical registration through exponentiated lie algebra (DARTEL)^41^. We applied a 6mm smoothing kernel. We entered the processed grey matter images in a two-sample t-test to compare grey matter volume (GMV) between dystonic tremor syndrome patients and controls. We added age and gender as covariates and corrected for brain volume differences by dividing voxel-by-voxel intensities by total intracranial volume (ICV).

### 2.8. Regions of interest

Given our a-priori hypothesis on involvement of both the cerebello-thalamo-cortical circuit and the basal ganglia (GPi) in dystonic tremor syndrome, we focused our analyses on these regions. The anatomical regions of interest we used were the cerebellum (using the SUIT toolbox), the GPi (taken from the Basal Ganglia Human Area Template toolbox^42^, Brodmann Area 4 (BA4, taken from the WFU PickAtlas^43,44^) and a combined mask of homologues of the thalamic VIM and VOp nuclei (posterior ventral lateral nucleus (VLp) and ventral lateral anterior (VLa) nucleus, taken from the Morel Atlas^45,46^). Here, we will follow the thalamic nomenclature by Hassler that is common among clinicians and neurosurgeons, with the ventral intermediate nucleus (VIM) and the ventral oralis posterior (VOp) nuclei being common targets for neurosurgical interventions for tremor^11,14^. Hassler’s nomenclature is an alternative for the nomenclature proposed by Jones^47^, where Hassler’s VIM is thought to correspond largely to Jones’s ventral lateral posterior (VLp) nucleus^48^, which receives input from the cerebellum. Hassler’s VOp is thought to correspond largely to Jones’ ventral lateral anterior (VLa) nucleus, which receives input from the pallidum. We used bilateral GPi, BA4 and thalamus ROIs in the structural analyses and unilateral left GPi, BA4 and thalamus ROIs in the functional analyses (matching the flipped functional contrast images).

### 2.9. Statistical analyses

For the analyses on task and tremor-related activity and the structural analyses, we assessed statistical significance with non-parametric permutation tests using the Threshold Free Cluster Enhancement (TFCE) Toolbox in SPM12 (http://dbm.neuro.uni-jena.de/tfce/) with 10,000 permutations. TFCE circumvents the problem of arbitrarily defined cluster forming thresholds while maintaining sensitivity with use of spatial neighbourhood information^49^. TFCE values represent the magnitude of evidence at each voxel combined with local spatial support. After TFCE, a voxel-level FWE-corrected p-value <0.05 was used as threshold for statistical significance inference. We increased this threshold to p<0.001 FWE-corrected for cerebellum-specific tremor-related activity and for hand lifting and lowering-related activity, to increase specificity in those analyses. With TFCE, we tested for tremor-related activity and grey matter volume differences in our regions of interest (ROI) and across the whole brain. The anatomy toolbox^50^ was used to anatomically localize clusters of activity.

For voxels with significant results, we related our findings with clinical tremor severity (sum score of TRS items of the most affected hand) and dystonia severity (Burke-Fahn-Marsden; BFM). For this, we averaged tremor-related activity and grey matter volume over significant voxels within ROIs using Marsbar^51^ (see Supplementary Table 2 for included ROIs). Given the predominant tremor-related activity in sensorimotor areas of the cerebellum^37^, we averaged over significant voxels in these areas for the cerebellum. We related tremor-related activity to clinical measures using Pearson or Spearman correlation coefficients, depending on normality of data distribution tested with Shapiro-Wilk Tests. For the structural analyses, we entered adjusted grey matter volumes (GMV/ICV), age and gender as independent variables into linear regression analyses with either clinical tremor severity or dystonia severity as dependent variable.

**Table 2.** Tremor related activity.

| Region | Side | Cluster size<br>(number of<br>voxels) | P<sub>FWE</sub> | TFCE-value | Coordinates peak voxels |  |  |
| --- | --- | --- | --- | --- | --- | --- | --- |
|  |  |  |  |  | x | y | z |
| Whole brain analysis |  |  |  |  |  |  |  |
| Cerebellum | L + R | 15519 | <0.001 | 3081 | -4 | -54 | -16 |
|  |  |  | <0.001 | 3022 | 8 | -62 | -26 |
|  |  |  | <0.001 | 3005 | 6 | -58 | -18 |
| Pallidum | L | 686 | 0.031 | 1234 | -16 | -8 | -4 |
|  |  |  | 0.032 | 1224 | -16 | -4 | 4 |
| Thalamus | L |  | 0.038 | 1158 | -20 | -16 | 6 |
|  |  |  | 0.038 | 1157 | -22 | -20 | 8 |
|  |  |  | 0.039 | 1139 | -16 | -6 | 12 |
| Pallidum | R |  | 0.042 | 1115 | 14 | -4 | -4 |
| Thalamus | R |  | 0.046 | 1080 | 12 | -8 | 6 |
|  |  |  | 0.046 | 1078 | 16 | -6 | 10 |
|  |  |  | 0.047 | 1072 | 18 | -14 | 10 |
| Frontal Pole | R | 1677 | 0.021 | 1387 | 14 | 60 | 30 |
|  |  |  | 0.022 | 1380 | 16 | 52 | 38 |
|  |  |  | 0.023 | 1363 | 20 | 56 | 24 |
|  | R | 48 | 0.038 | 1157 | 38 | 50 | -18 |
|  | R | 2 | 0.049 | 1047 | 38 | 48 | -8 |
| Frontal Pole | L | 6 | 0.049 | 1055 | -18 | 52 | 16 |
| Brainstem | R | 74 | 0.041 | 1118 | 12 | -16 | -16 |
| Temporal Pole | L | 113 | 0.026 | 1314 | -54 | 2 | -38 |
| Cerebellum specific analysis |  |  |  |  |  |  |  |
| Right Lobule I<sub>IV</sub> / V | R | 31149 | <0.001 | 6772 | 8 | -61 | -26 |
|  |  |  | <0.001 | 6687 | 7 | -57 | -19 |
|  |  |  | <0.001 | 6184 | 20 | -53 | -23 |
| Vermis VI / VIIa | medial |  | <0.001 | 6568 | 0 | -62 | -29 |
|  |  |  | <0.001 | 6188 | 2 | -65 | -34 |
|  |  |  | <0.001 | 6186 | 5 | -66 | -39 |
| Left Lobule I<sub>IV</sub> / V | L |  | <0.001 | 6703 | -4 | -54 | -13 |
|  |  |  | <0.001 | 6696 | -5 | -58 | -16 |
|  |  |  | <0.001 | 6677 | -4 | -62 | -16 |
|  |  |  | <0.001 | 6076 | -5 | -47 | -17 |
| Right_Crus I & II | R | 570 | 0.001 | 3811 | 43 | -57 | -45 |
|  |  |  | 0.001 | 3796 | 44 | -62 | -35 |
|  |  |  | 0.001 | 3670 | 37 | -63 | -41 |

For the thalamus ROI, which showed both tremor related activity and increased GMV in patients, we tested whether these two measures correlated. We extracted adjusted GMV from the most tremulous side and entered this, again with age and gender, in a linear regression analysis with tremor-related activity (average beta over thalamus ROI) as dependent variable. Finally, we correlated tremor-related activity between the different regions of interest, to explore whether a trade-off between brain regions was present (e.g. relatively more cerebellum involvement in some patients and more pallidum involvement in others). We used a statistical significance threshold of p<0.05 for all correlation analyses.

## 3. Results

Characteristics of dystonic tremor syndrome patients are presented in Table 1. Posture holding significantly increased tremor power (posture holding vs. non-posture holding, log tremor power 9.0 ± 1.2 versus 6.2 ± 0.6, *t*_*(26)*_ = 11.02, p < 0.001, Supplementary Fig. 1). The majority of patients (85%, *n*=23) were diagnosed as dystonic tremor (dystonia in tremulous hand/arm), and 15% (*n*=4) as tremor associated with dystonia (dystonia in body part other than tremulous hand/arm). Tremor was judged to be slightly irregular in 59% of patients, asymmetrical in 74%, and posture-dependent in 67%. None of the patients had myoclonus.

### 3.1. Tremor-related brain activity

We observed significant tremor-related activity in the cerebellum, pallidum and thalamus, bilateral frontal pole, and small clusters in the brainstem and temporal pole (whole brain analysis, Fig. 2, Table 2). ROI analyses revealed that tremor-related activity was most pronounced in sensorimotor regions of the cerebellum, i.e. right lobule I-IV and V, right crus I&II, vermis VI and VIIIa, and left lobules I-IV and V^37^ (Table 2). When testing in our other three ROIs, we found significant tremor-related activity in the left GPi (15 voxels, MNI local maximum [-18 -8 -4], *TFCE* = 71.35, *p(fwe)* = 0.005), the left thalamus (88 voxels, MNI local maximum [-18 - 16 12], *TFCE* = 77.05, *p(fwe)* = 0.012) and the left BA4 (hand area of the primary motor cortex, 130 voxels, MNI local maximum [-36 -26 56], *TFCE* = 151.88, *p(fwe)* = 0.002), see Fig. 2B. Within the thalamus, tremor-related activity could be localized to both the VIM (27.63% of the dorsal VLp and 31.22% ventral VLp were activated) and the VOp (69.91% of the VLa activated). Tremor-related activity did not differ between the thalamic nuclei (mean beta over whole VLa = 0.045 ± 0.096, VLpv = 0.040 ± 0.096 and VLpd = 0.043 ± 0.107, one-way repeated measures ANOVA *F(2,52)* = 0.144, p = 0.866). Tremor-related activity did not correlate with clinical tremor severity or dystonia severity in any of the ROIs (Supplementary Table 2). Tremor-related activity in the cerebellum showed a significant positive correlation with tremor-related activity in the GPi (Fig. 2C and Table 3).

**Table 3.** Correlation coefficients of tremor-related activity between ROIs.

|  | Cerebellum | GPI | VLp | BA4 |
| --- | --- | --- | --- | --- |
| **Cerebellum** | - |  |  |  |
| **GPI** | 0.546 (0.003) | - |  |  |
| **Thalamus** | 0.820 (<0.001) | 0.679 (<0.001) | - |  |
| **BA4** | 0.778 (<0.001) | 0.515 (0.006) | 0.687 (<0.001) | - |

Values displayed as: Correlation coefficient (p-value)
Spearman's Rho (Thalamus) or Pearson's R (other ROIs).

**Figure 2.**
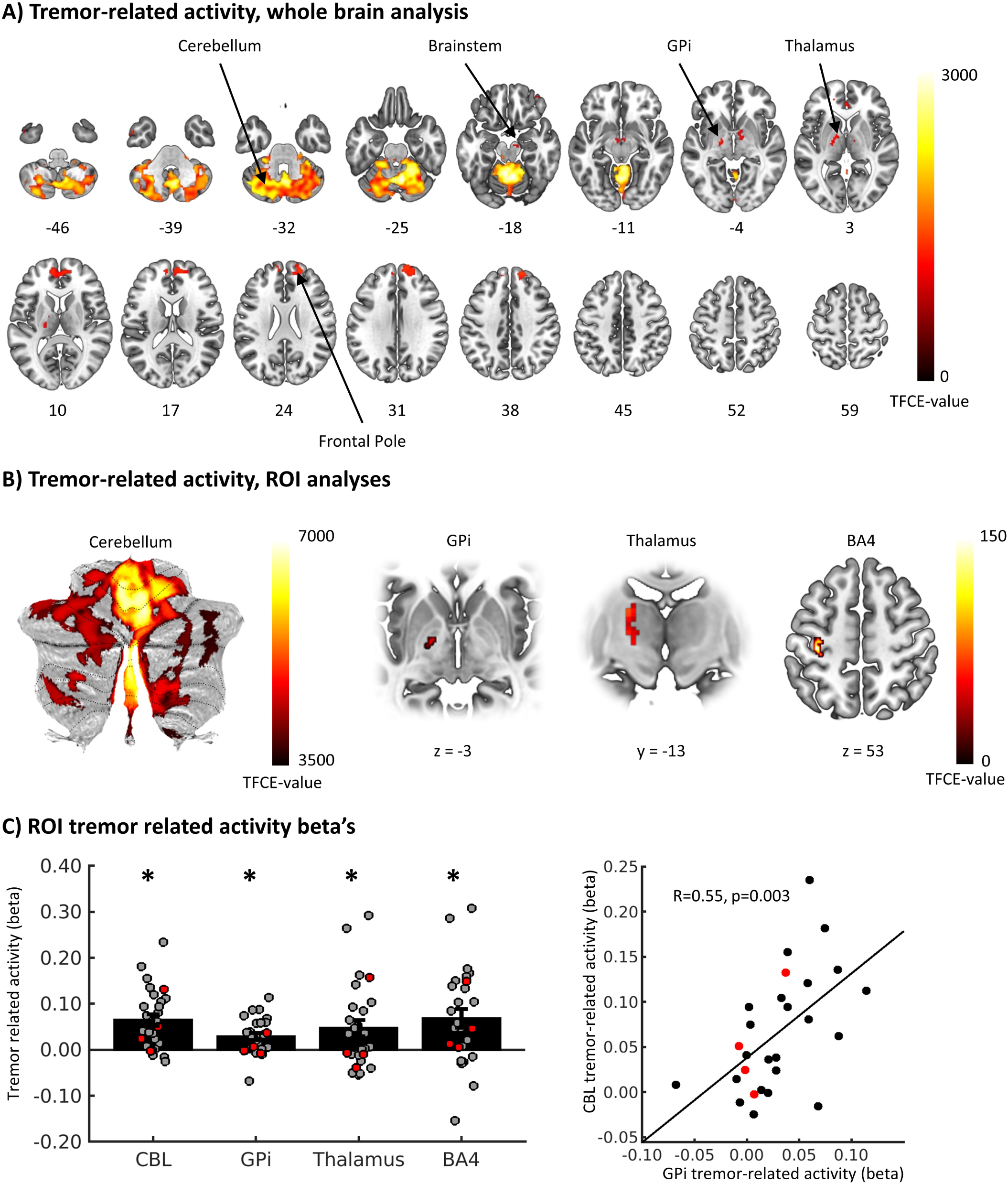
Tremor-related activity. **(A)** Brain regions with significant tremor-related activity in the whole brain analysis. Values represent z-axis MNI coordinates. **(B)** Voxels with significant tremor related activity in the region of interest analyses. **(C)** Tremor-related activity averaged over significant voxels in the somatomotor regions of the cerebellum, the GPi, VIM and BA4 (left plot). Bars represent mean beta values (± SEM), dots represent individual beta’s and asterisks indicate statistical significance. The right plot shows a positive correlation between cerebellar and GPi tremor related activity. Patients with dystonic tremor are displayed in grey/back dots, patient with tremor associated with dystonia in red dots. GPi = internal globus pallidus, TFCE = Threshold free cluster enhancement, ROI = Region of interest, VIM = ventral intermediate nucleus, BA4 = Brodmann Area 4, CBL = Cerebellum.

### 3.2. Hand-lifting related brain activity

By design, tremor-related activity was specific to fluctuations in tremor power and not associated with hand lifting, posture holding, or hand lowering. Hand lifting and hand lowering were associated with a specific pattern of activity in the contralateral sensorimotor cortex and ipsilateral cerebellum (laterality related to the used hand), as well as a more widespread motor circuit during hand lowering. Furthermore, stable posture holding, defined as holding the hand in the predefined posture, was related to brain activity in the ipsilateral cerebellum. See Supplementary Table 1 and Supplementary Fig. 2 for more (statistical) details.

### 3.3. Effective connectivity

Bayesian model selection revealed that during hand lifting, which coincides with the onset of tremor, network activity most likely starts in BA4, rather than in the cerebellum, thalamus or GPi (highest probability for an input [DCM.C] onto BA4: expected posterior probability = 0.43, exceedance probability = 0.83, Fig. 3a). Furthermore, Bayesian model averaging showed that connectivity from the cerebellum to thalamus (*DCM*.*B* = 0.60 ± 0.08 Hz, *t(26)* = 2.92, *p(FDR-corrected*)=0.039) and the thalamus inhibitory self-connection (*DCM*.*B* = 1.35 ± 0.09 Hz, *t(26)* = 4.12, p(FDR-corrected) = 0.004) were positively modulated by tremor amplitude fluctuations (Fig. 3 B,C). These effects were not observed for connectivity between the GPi and thalamus or any other region (*p(FDR-corrected)* > 0.05). This suggests that individual fluctuations in tremor amplitude are specifically driven by cerebello-thalamic connectivity. These connectivity parameters were not correlated with clinical tremor or dystonia severity (Supplementary Table 2).

**Figure 3.**
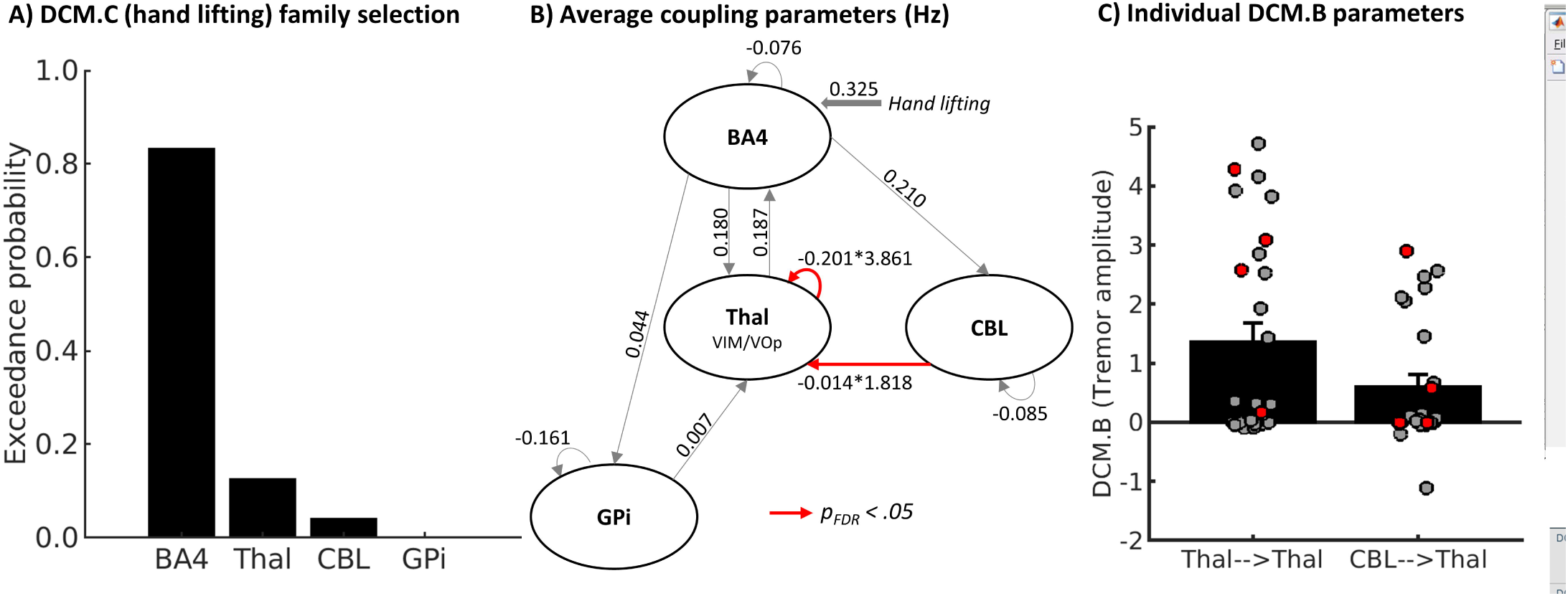
Results of dynamic causal modelling. **(A)** Bayesian model selection comparing families of models with input of hand lifting (DCM.C) on BA4, Thalamus, Cerebellum or GPi **(B)** Mean coupling parameters resulting from Bayesian model averaging over all models with input of hand lifting on BA4. Intrinsic connectivity parameters (DCM.A) are displayed for all connections. For connection with significant modulation of tremor amplitude (Cerebellum to thalamus and thalamus self-connection) the net coupling parameters are calculated according to DCM.A*exp(DCM.B). **(C)** Individual DCM.B parameters for connections with significant modulation of tremor amplitude. Patients with dystonic tremor are displayed in grey/back dots, patient with tremor associated with dystonia in red dots. BA4 = Brodmann Area 4, Thal = Thalamus, CBL = Cerebellum, GPi = internal globus pallidum, FDR = False discovery rate.

### 3.4. Grey matter volume

ROI analyses showed significantly increased grey matter volume in patients versus controls in BA4 (two bilateral medial clusters and a left lateral cluster) and in the left thalamus (Table 4, Fig. 4). Within the thalamus, increased grey matter volume was localised in VIM (49.74% of the dorsal VLp and 24.50% of the ventral VLP showed increased grey matter volume) and the VOp (28.84% of the VLa with increased grey matter volume). Within the thalamus, there was an overlap between voxels showing tremor-related activity and voxels showing increased grey mater volume. In contrast, in BA4 the region showing increased grey matter was located more medially than the region showing tremor-related activity, without overlap. Grey matter volume in these clusters did not correlate with clinical tremor severity or dystonia severity. In the thalamus, grey matter volume did not correlate with tremor-related activity. Whole brain analyses revealed no other regions showing significant group differences.

**Table 4.**
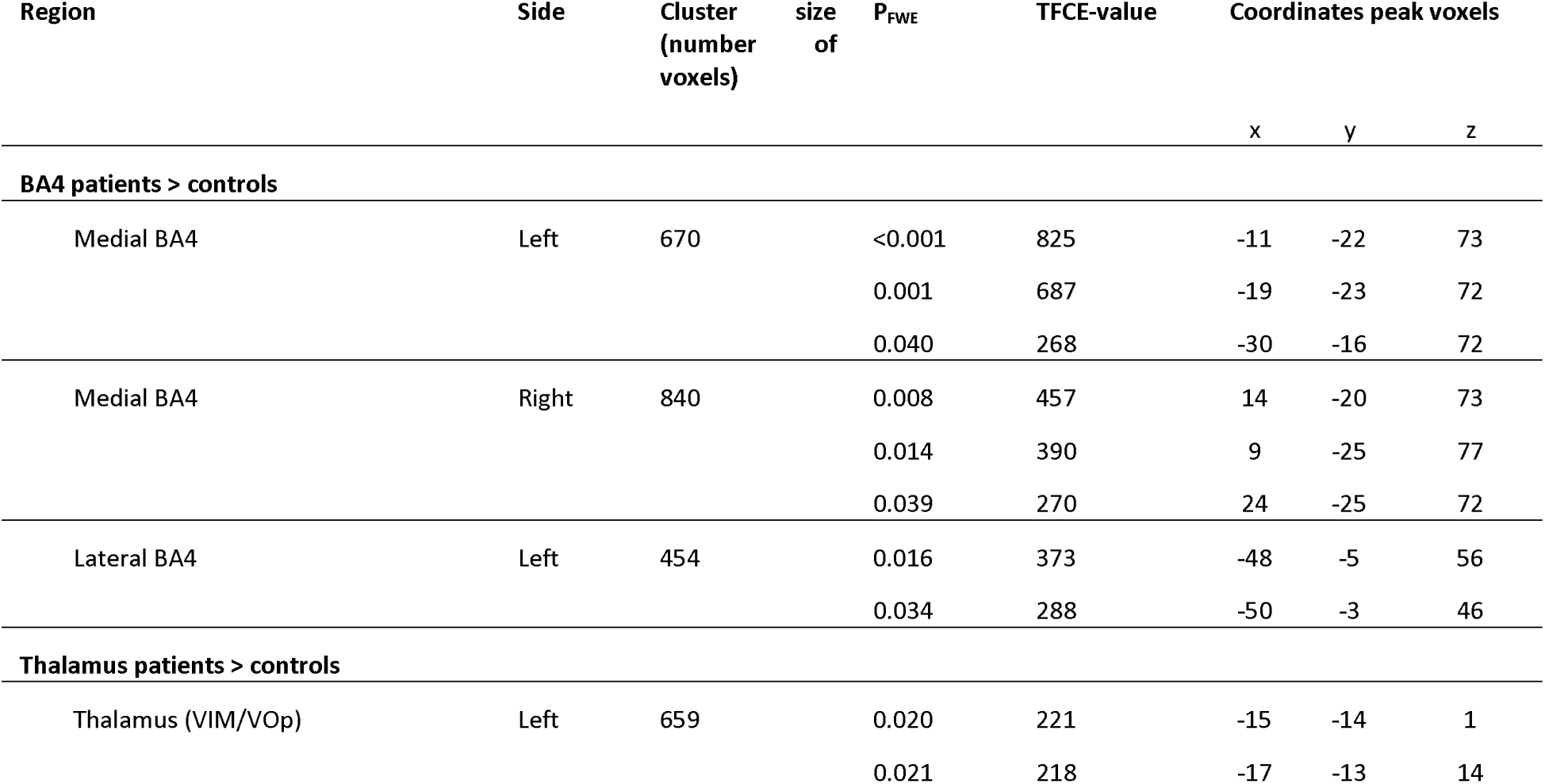
Grey matter volume differences between dystonic tremor patients and healthy controls, region of interest analyses.

**Figure 4.**
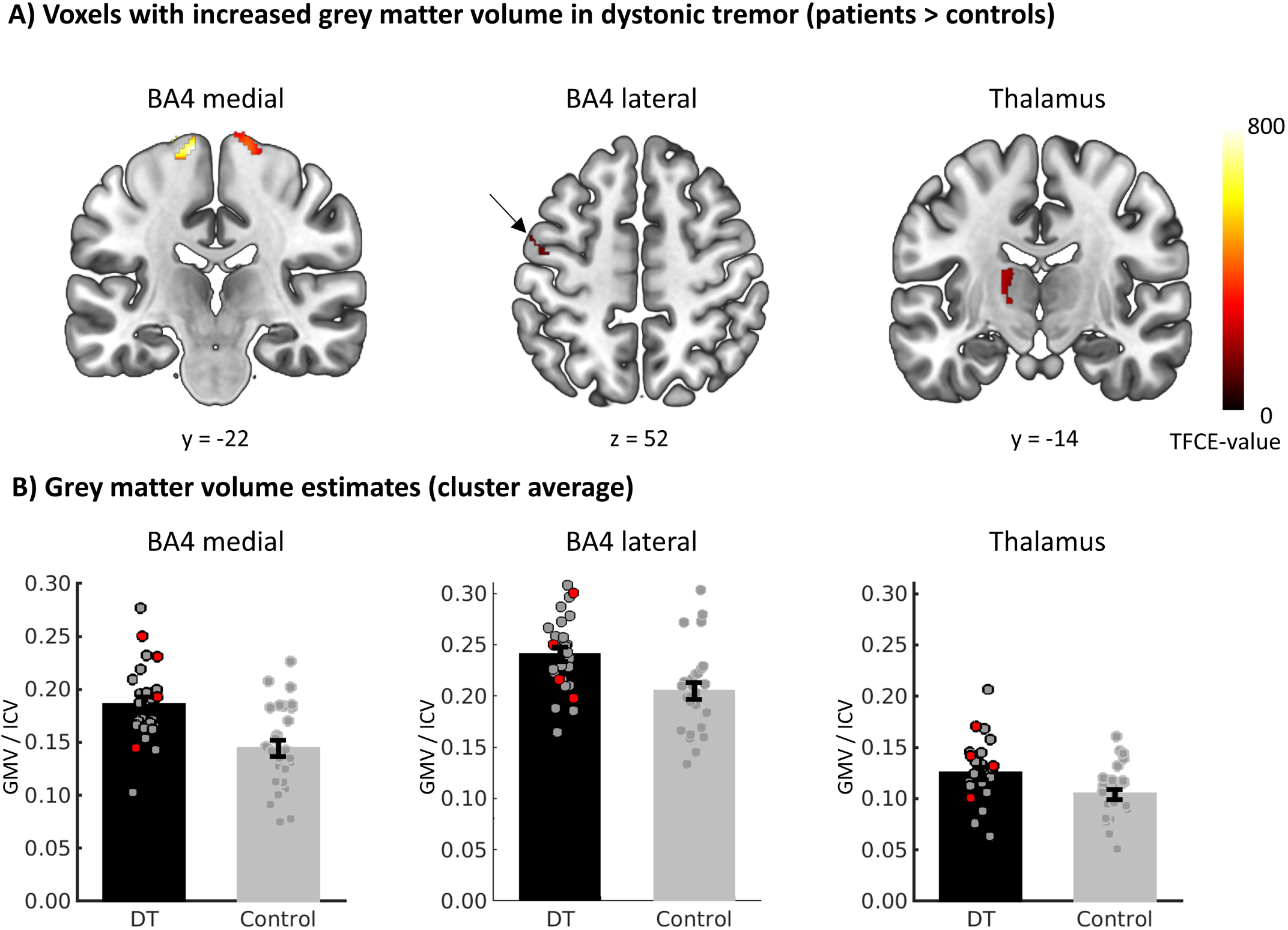
Grey matter volume differences between dystonic tremor patients and healthy controls. **(A)** Voxels with significantly higher grey matter volume in dystonic tremor patients compared to controls in region of interest analyses. **(B)** Grey matter volume estimates, adjusted for intracranial volume and averaged over significant voxels displayed in A. Bars represent mean (± SEM) and dots represent individual grey matter volumes. Patients with dystonic tremor are displayed in grey/back dots, patient with tremor associated with dystonia in red dots. BA4 = Brodmann Area 4, VIM = ventral intermediate nucleus, TFCE = Threshold free cluster enhancement. GMV = grey matter volume, ICV = Intracranial volume.

## 4. Discussion

We investigated cerebral activity associated with tremor in dystonia. To this end, we disentangled cerebral activity specifically related to tremor fluctuations from activity related to (voluntary) posture holding. There are three main findings. First, tremor-related activity was present in both the cerebello-thalamic-cortical circuit (motor regions of the cerebellum, thalamus (VIM), hand area in the primary motor cortex), and in the basal ganglia (GPi, as well as the pallidal recipient thalamic nucleus VOp). Second, effective connectivity from cerebellum to thalamus and thalamus self-inhibition were associated with tremor amplitude fluctuations. Third, grey matter volume was increased in patients compared to controls, in the same thalamic region showing tremor-related activity (both VIM and VOp), and in two regions within the primary motor cortex where we did not observe tremor-related activity (bilateral medial and left lateral BA4). Below we discuss how these findings help to understand the complex pathophysiology underlying dystonic tremor syndromes.

### 4.1. Dystonic tremor syndrome: the role of the cerebello-thalamo-cortical circuit

We confirmed our hypothesis that both the cerebello-thalamic circuit and the basal ganglia are involved in dystonic tremor syndrome, although the extent of tremor-related activity in the cerebellum was much larger than in the other regions. This fits with behavioural findings suggesting that the cerebellum plays a key role in the pathophysiology of dystonic tremor syndrome. Specifically, the temporal prediction of motion perception and classical eye blink conditioning, both markers of cerebellar dysfunction, are impaired in dystonia patients with tremor versus without tremor^23,58^. Furthermore, in a mouse model of dystonia, blocking of glutamatergic olivocerebellar signalling caused a limb tremor^59^. The tremor-related activity we observed was specific to fluctuations in tremor, and therefore goes beyond a more general “trait” of dystonia. Nevertheless, the role of the cerebellum in dystonic tremor syndrome, as shown here, aligns with more general ideas that the cerebellum plays a critical role in the pathophysiology of dystonia^60^. This is evidenced by functional and structural alterations in the cerebellum and its connections to thalamus and primary motor cortex^61^. Here we did not find evidence for cerebellar atrophy in dystonia patients, in line with previous work^62^. This suggests that the role of the cerebellum in dystonic tremor syndrome is associated with functional rather than structural changes. A possible mechanism underlying tremor-related activity in the cerebellum could be a loss of inhibition due to impaired GABAergic transmission. More specifically, a PET study has shown that focal hand dystonia patients showed a lower flumazenil binding potential than controls in the vermis VI of the ipsilateral cerebellum and the contralateral sensorimotor cortex (with respect to the dystonic arm), while there were no differences in the striatum^62^.

The functional role of the cerebellum in dystonic tremor syndrome is further qualified by our finding that effective connectivity from cerebellum to thalamus was modulated by tremor amplitude fluctuations. Specifically, we found a negative endogenous connection between cerebellum and thalamus (the DCM.A parameter in Fig. 3), which was positively modulated by intra-individual fluctuations in tremor power (the DCM.B parameter in Fig. 3). This is consistent with findings in a DCM study in essential tremor, which showed the same combination of findings^17^. The negative endogenous connection fits the functional anatomy of the cerebello-thalamic pathway, which consists of two projections: an inhibitory connection between Purkinje cells in the cerebellar cortex and the deep cerebellar nuclei, and an excitatory connection between the deep cerebellar nuclei and thalamic nuclei (VIM)^63^. The positive modulation of this endogenous connection by tremor amplitude, together with widespread tremor-related activity in the cerebellar cortex, suggests that abnormal activity in the cerebellar cortex drives tremor in dystonia through the thalamus. We could not empirically test whether tremor-related activity was indeed initiated in the cerebellum, since the onset of tremor coincides with the onset of hand-movement (DCM.C parameter Fig. 3, which had its input on BA4). However, the involvement of cerebellum-thalamus connectivity in dystonic tremor syndrome aligns with previous studies. First, DBS for dystonic tremor was successful when targeted at the anatomical connection between cerebellum and thalamus, the dentato-rubro-thalamic tract^14,64^. Second, functional coupling between cerebellum and thalamus was lower in dystonic tremor patients compared to essential tremor patients and controls during a grip-force task^15^. Third, a recent transcranial magnetic stimulation (TMS) study showed that cerebello-cortical inhibition was reduced in patients with dystonic tremor as compared to controls and essential tremor patients^65^. An unexpected finding is that fluctuations in tremor power were associated with increased (rather than reduced) inhibitory self-connectivity of the thalamus. In Parkinson’s disease tremor, dopaminergic medication (which reduces tremor) has previously been shown to specifically increase thalamus inhibition^38,66^. In the context of a cerebellar drive, the modulation of tremor amplitude on thalamic self-inhibition can be interpreted as an endogenous attempt to suppress tremor-related activity that arises in the cerebellum. Such self-regulatory mechanisms are thought to be necessary for protecting the brain against non-linear increases in neural activity, which would otherwise result in epileptic seizures. VIM-DBS may thus reduce dystonic tremor syndromes (as well as other tremors) by interfering with cerebello-thalamic transmission of tremor oscillations^67^, and by potentiating thalamic self-inhibition^68^.

Regional changes in grey matter volume in other nodes of the cerebello-thalamo-cortical circuit in dystonia may further contribute to tremor pathophysiology. That is, although we found no evidence of structural cerebellar alterations in the patients, we did observe increased grey matter volume in the thalamus (in both VIM and VOp) in patients compared to controls. This has previously also been observed in patients with Parkinson’s resting tremor^69^, underlining the more general role of the thalamus in tremor. Given that the increase in grey matter in the thalamus overlapped with voxels showing tremor-related activity, it is very likely that the structural changes are related to tremor. Whether these structural changes cause tremor or result from prolonged tremor-related activity^70^ remains unknown. In addition, we also found increased grey matter volume in BA4, which aligns with a previous report on cortical thickening in the sensorimotor cortex in dystonic tremor patients compared to controls^22^. The grey matter changes in BA4 did not overlap with tremor-related activity in BA4 and did not correlate with clinical dystonia or tremor severity scores, so it remains unclear whether this finding is related to the dystonia, to tremor, or both.

### 4.2. Dystonic tremor syndrome: the role of the basal ganglia

Besides tremor-related activity in all nodes of the cerebello-thalamo-cortical network, we also found tremor-related responses in the basal ganglia (GPi), and the frontal pole. This underlines that the pathophysiology of dystonic tremor syndrome is not restricted to a single brain region or even a single network, similar to dystonia itself^60,61^. The tremor-related activity in the GPi and its thalamic relay nucleus (the VOp) suggests involvement of the pallido-thalamic pathway in dystonic tremor syndromes.

This is further supported by a study that reported a sweet spot for DBS in dystonic tremor on the border between VIM and VOp^14^. A clinically relevant question is how tremor-related activity in the cerebello-thalamic and the pallido-thalamic pathways relate to each other. The VIM and GPi are two popular targets for tremor suppression with stereotactic surgery^7,10-13^, but treatment is not always effective in reducing dystonic tremor syndrome. For instance, in one study, three out of six dystonic tremor patients first treated with VIM-DBS were subsequently treated with GPi-DBS due to lack of effect^71^. This may suggest that there is a trade-off between the involvement of the two circuits, where some patients have a “cerebellar-type” tremor and others a “basal ganglia-type” tremor^8,9^. Here we found that, across the entire sample, the degree of tremor-related activity in GPi and cerebellum was positively correlated. This goes against the idea of a trade-off, but rather suggests that the basal ganglia and cerebellum act in concert. Specifically, tremor-related activity may be propagated from the cerebellum to the basal ganglia (or vice versa) via reciprocal subcortical anatomical connections between these two structures^72^, or through the motor cortex – as is the case in Parkinson’s resting tremor^31^. Indeed, in patients with cervical dystonia without tremor, abnormal functional connectivity between basal ganglia and cerebellum has been found^73^.

We did not find a relationship between the presence or absence of clinical tremor characteristics (such as regularity of the tremor) and the magnitude of tremor-related activity in GPi versus the cerebellum. Support for such differential pathophysiology between tremor phenotypes comes from pallidal single neuron recordings during DBS^74^. In pure dystonia and dystonia with jerky head tremor, relatively more burst cells were present, while relatively more pause cells were present in dystonia with sinusoidal head tremor. This suggest that jerky head tremor relies on pallidal alterations, while sinusoidal head tremor relies on other, possibly cerebellar, alterations. Whether this also holds for hand tremor, and which cell types contributed to the BOLD signal we observed, remains unknown. Finally, we also observed tremor-related activity in the frontal pole. This effect might be related to increased cognitive monitoring or arousal during episodes of high-tremor^26^. Future studies with a wider spectrum of tremor phenotypes, including essential tremor, essential tremor plus and dystonic tremor syndrome patients, may further investigate whether there is a gradient between cerebellar and pallidal contributions to tremor depending on the phenotype. Furthermore, studies investigating whether the degree of cerebellar versus pallidal tremor-related activity may predict DBS treatment effects are worthwhile.

### 4.3. Interpretational issues

First, we did not include a control group for our functional MRI analyses, which raises the question to what extent the findings are specific to dystonic tremor syndrome. We purposely did not include a healthy control group that mimicked tremor, because mimicked (voluntary) and pathological (involuntary) tremor differ in many ways, undermining the value of a direct comparison. For instance, voluntary movements involve motor planning (inverse models) and altered weighing of somatosensory feedback (forward modelling), as compared to involuntary movements^75^. Furthermore, mimicked tremor phenotypically differs from pathological tremor in fundamental ways: even in subjects who were explicitly instructed to mimic essential tremor as well as possible, voluntary tremor had a lower frequency and larger wrist extension-flexion movement compared to essential tremor^16^. Second, we were unable to show direct correlations between tremor-related activity and clinical tremor or dystonia severity. This raises the question whether the cerebral activity we report is clinically meaningful. On the other hand, it may also suggest that the findings we report are shared across a wide range of dystonia patients with tremor, while larger groups are necessary to detect cerebral sources of inter-individual variability. A third issue we need to consider concerns the included population. We combined patients with dystonic tremor (dystonia in tremulous hand) and with tremor associated with dystonia (dystonia in body part other than tremulous hand). Dystonic tremor was more prevalent than tremor associated with dystonia (85% versus 15% of included patients). This matches the prevalence in a large cohort-study in which 715 patients with dystonic tremor syndrome were included. Of these 715 patients, tremor was classified as dystonic tremor in 614 patients (86%) and as associated with dystonia in 101 patients (14%)^3^. Given that recent findings have indicated pathophysiological differences between DT and TAWD^65^, we considered the possibility that the 4 TAWD patients in our sample (indicated in red in the figures) may have introduced considerable inter-individual variability. Hence, we repeated our main analyses by including dystonic tremor syndrome subtype (DT versus TAWD) as a covariate. This did not change any of our findings (Supplementary Table 3). Exclusion of the 4 TAWD patients also left our main findings intact (Supplementary Table 3). Although this study was not designed (or powered) to investigate cerebral differences between dystonic tremor syndrome subtypes, the analyses outlined above indicate that the cerebral effects reported here are shared across dystonic tremor syndrome phenotypes.

## 5. Conclusions

The findings show that both the cerebello-thalamo-cortical circuit and the basal ganglia are involved in the pathophysiology of dystonic tremor syndromes. Specifically, tremor-related activity was present in the cerebellum, the thalamus (VIM and VOp), the primary motor cortex and the GPi. Given the predominant tremor-related activity in the cerebellum, and altered cerebello-thalamic effective connectivity, we propose that tremor in dystonia may be driven by cerebellar dysfunction. Grey matter hypertrophy of other nodes of the cerebello-thalamo-cortical circuit, i.e. the VIM, VOp and motor cortex, may further contribute to an imbalance in this network.

## Data Availability

All data produced in the present study are available upon reasonable request to the authors

## 6. Acknowledgements

We thank all participants and their spouses for investing their time and effort in this study. Also, we thank everybody that helped with recruitment of participants. We acknowledge the funding from the Netherlands Brain Foundation and the Benny Vleerlaag fonds.

