## Supplementary for "Cerebello-thalamic activity drives an abnormal motor network into dystonic tremor"

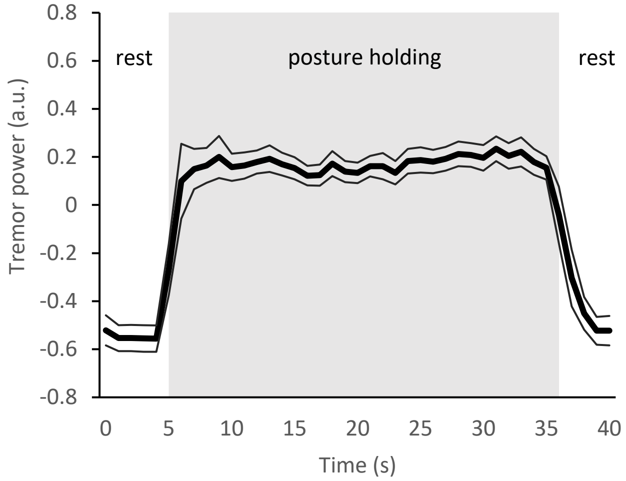


**Supplementary figure 1. Tremor power during rest and posture holding blocks**

Tremor power over time at peak tremor frequency averaged over 20 blocks within participants and then over participants. Mean tremor power (thick black line) ± SEM (thin black lines). a.u. = arbitrary units.

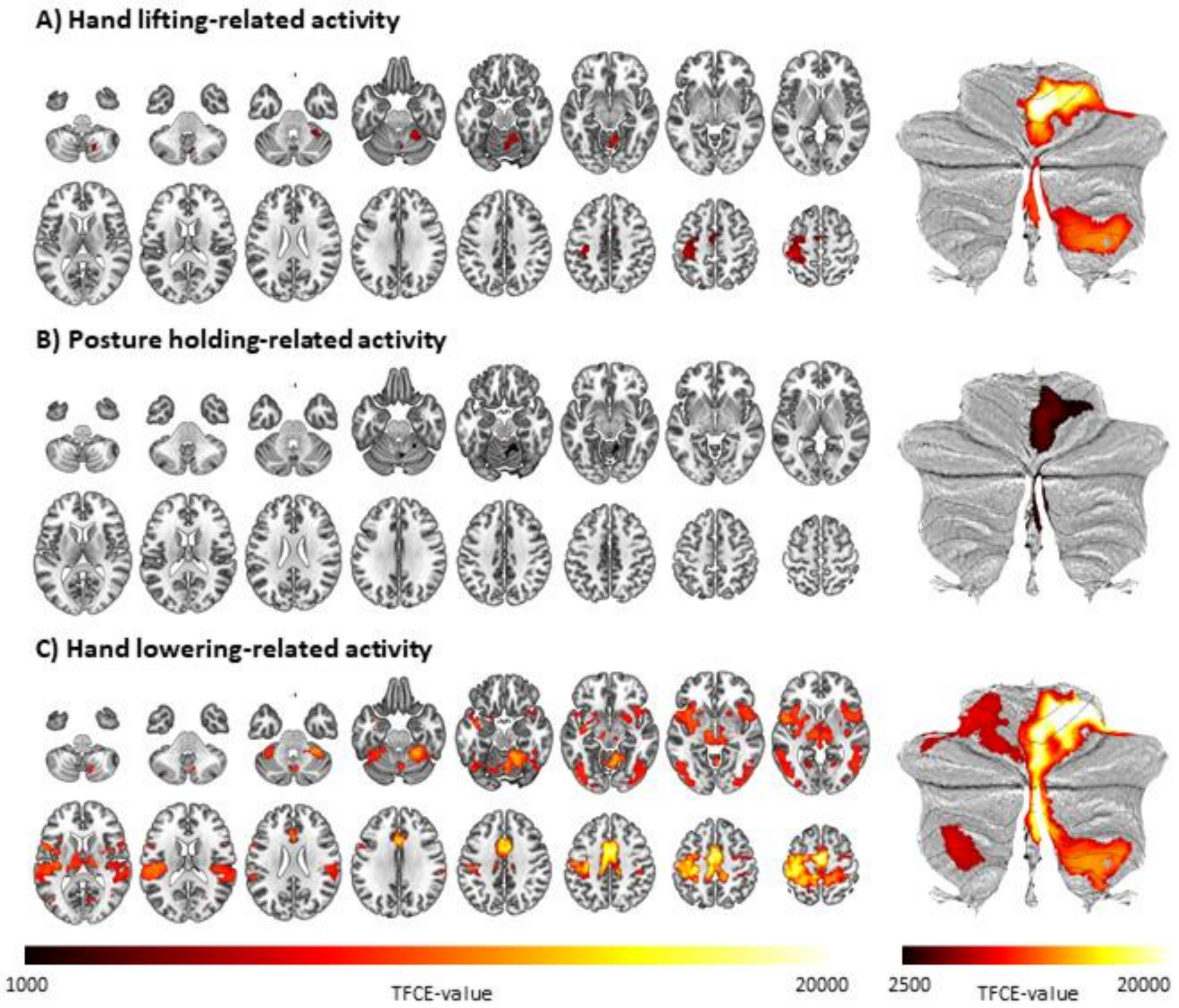


**Supplementary figure 2. Posturing-related brain activity**

Voxels with significant hand lifting (A), posture holding (B) and hand lowering (C) related brain activity. Results from whole brain analysis are displayed on the left, from cerebellum specific analyses on the right. TFCE = Threshold free cluster enhancement.

| Supplementary Table 1. Hand lifting, posture holding and lowering-related brain activity | | | | | | | |
|---|---|---|---|---|---|---|---|
| **Region** | | **Cluster size (number of voxels)** | $P_{FWE}$ | **TFCE-value** | **Coordinates peak voxels** | | |
| | | | | | x | y | z |
| **Hand lifting-related activity** | | | | | | | |
| Pre and Postcentral gyri (hand sensorimotor area) | | 2016 | <0.001 | 8135 | 6 | -58 | -12 |
| | | | <0.001 | 7441 | 12 | -54 | -14 |
| | | | <0.001 | 7298 | 10 | -50 | -14 |
| | | | <0.001 | 6872 | 20 | -56 | -58 |
| | | | <0.001 | 6564 | 28 | -46 | -28 |
| | | | <0.001 | 6435 | 22 | -54 | -22 |
| | | | <0.001 | 6421 | 22 | -50 | -24 |
| | | | <0.001 | 6345 | 28 | -56 | -58 |
| | | | <0.001 | 5840 | 14 | -62 | -58 |
| | | | <0.001 | 5693 | 6 | -66 | -22 |
| | | | <0.001 | 5686 | 4 | -68 | -18 |
| | | | <0.001 | 4981 | 28 | -46 | -54 |
| | | | <0.001 | 4270 | 6 | -68 | -38 |
| | | | <0.001 | 4118 | 6 | -72 | -46 |
| | | | <0.001 | 4118 | 12 | -70 | -42 |
| | | | 0.001 | 3672 | 20 | -66 | -22 |
| Cerebellum | | 3292 | <0.001 | 7424 | -42 | -34 | 60 |
| | | | <0.001 | 7413 | -38 | -34 | 68 |
| | | | <0.001 | 7381 | -36 | -32 | 60 |
| | | | <0.001 | 7285 | -32 | -30 | 72 |
| | | | <0.001 | 7176 | -40 | -24 | 70 |
| | | | <0.001 | 6580 | -32 | -26 | 60 |
| | | | <0.001 | 6570 | -34 | -28 | 46 |
| | | | <0.001 | 6542 | -26 | -48 | 62 |
| | | | <0.001 | 6521 | -42 | -28 | 50 |
| | | | <0.001 | 6518 | -28 | -26 | 48 |
| | | | <0.001 | 6484 | -38 | -32 | 52 |
| | | | <0.001 | 6018 | -18 | -48 | 68 |
| | | | <0.001 | 5800 | -40 | -22 | 54 |
| | | | <0.001 | 5800 | -26 | -38 | 70 |
| | | | <0.001 | 5701 | -24 | -10 | 64 |
| | | | <0.001 | 5657 | -28 | -14 | 64 |
| Juxtapositional Lobule / anterior division Cingulate Gyrus (SMA / preSMA) | | 565 | <0.001 | 4412 | -6 | -6 | 50 |
| | | | <0.001 | 4348 | -8 | -14 | 50 |
| | | | <0.001 | 4119 | 6 | -2 | 52 |
| | | | <0.001 | 4112 | 6 | 0 | 48 |
| | | | <0.001 | 4112 | 6 | -4 | 56 |

| | | | | | | | |
|---|---|---|---|---|---|---|---|
| | | | <0.001 | 4066 | 8 | 2 | 44 |
| | | | 0.001 | 3796 | -6 | -24 | 52 |
| | | | 0.001 | 3791 | -6 | -2 | 40 |
| | | | 0.001 | 3668 | -12 | 4 | 42 |
| **Posture holding-related activity** | | | | | | | |
| Cerebellum | | 921 | 0.004 | 1712 | 6 | -60 | -22 |
| | | | 0.005 | 1674 | 6 | -58 | -12 |
| | | | 0.005 | 1623 | 8 | -48 | -14 |
| | | | 0.006 | 1613 | 12 | -50 | -16 |
| | | | 0.013 | 1380 | 20 | -54 | -18 |
| | | | 0.014 | 1352 | 26 | -58 | -60 |
| | | | 0.015 | 1336 | 6 | -68 | -36 |
| | | | 0.016 | 1318 | 20 | -62 | -50 |
| | | | 0.016 | 1313 | 16 | -64 | -46 |
| | | | 0.016 | 1303 | 22 | -62 | -54 |
| | | | 0.027 | 1154 | 24 | -54 | -24 |
| | | | 0.028 | 1153 | 22 | -48 | -26 |
| | | | 0.031 | 1119 | 14 | -64 | -58 |
| | | | 0.035 | 1084 | 28 | -48 | -28 |
| **Onset-related activity** | | | | | | | |
| Sensorimotor network | | 41366 | <0.001 | 19408 | -2 | -10 | 58 |
| | | | <0.001 | 19332 | -2 | -10 | 64 |
| | | | <0.001 | 18832 | -4 | -10 | 52 |
| | | | <0.001 | 18712 | 4 | -6 | 58 |
| | | | <0.001 | 18285 | -2 | -4 | 50 |
| | | | <0.001 | 18243 | -4 | -2 | 46 |
| | | | <0.001 | 18087 | 2 | -2 | 50 |
| | | | <0.001 | 17986 | 4 | 0 | 46 |
| | | | <0.001 | 17558 | -38 | -12 | 66 |
| | | | <0.001 | 17387 | -36 | -30 | 62 |
| | | | <0.001 | 17296 | -42 | -30 | 66 |
| | | | <0.001 | 17227 | -36 | -22 | 60 |
| | | | <0.001 | 17182 | 2 | -4 | 70 |
| | | | <0.001 | 16961 | 6 | -2 | 64 |
| | | | <0.001 | 16791 | 6 | -14 | 58 |
| | | | <0.001 | 16631 | 6 | -12 | 46 |
| Insula (Area Ig1) | | 1 | 0.001 | 6425 | 32 | -30 | 10 |

**Supplementary Table 2. Correlations between functional MRI outcomes and clinical measures**

| | Tremor severity (TRS-ma) | Dystonia severity (BFM) |
|---|---|---|
| **Tremor related activity** | | |
| GPi | -0.119 (0.553)[1] | -0.098 (0.626)[2] |
| CBL | -0.054 (0.790)[1] | -0.235 (0.237)[2] |
| Thalamus | -0.129 (0.521)[2] | -0.195 (0.330)[2] |
| BA4 | -0.017 (0.934)[1] | -0.248 (0.212)[2] |
| **Effective connectivity DCM.B** | | |
| Cerebellum to thalamus | 0.277 (0.162)[2] | 0.162 (0.420)[2] |
| Thalamus self-connection | 0.086 (0.671)[2] | -0.069 (0.733)[2] |

[1]Pearson's R

[2]Spearman's Rho

**Supplementary Table 3. Tremor related activity, post-hoc whole brain analyses to determine the effect of tremor type: tremor associated with dystonia (N=4) vs. dystonic tremor (N=23)**

| Region | Side | Cluster size (number of voxels) | $P_{FWE}$ | TFCE-value | Coordinates peak voxels | | |
|---|---|---|---|---|---|---|---|
| | | | | | x | y | z |
| **Tremor associated with dystonia as covariate (N=27)** | | | | | | | |
| Cerebellum | L+R | 14095 | 0.001 | 2968 | -4 | -54 | -16 |
| | | | 0.001 | 2918 | 8 | -62 | -26 |
| | | | 0.001 | 2895 | 6 | -58 | -18 |
| Pallidum | L | 202 | 0.037 | 1188 | -16 | -8 | -6 |
| | | | 0.038 | 1179 | -16 | -4 | 4 |
| Thalamus L | L | | 0.044 | 1111 | -20 | -16 | 6 |
| Pallidum | R | 14 | 0.048 | 1073 | 14 | -4 | -4 |
| Frontal Pole | R | 1301 | 0.026 | 1341 | 12 | 60 | 32 |
| | | | 0.027 | 1326 | 16 | 52 | 38 |
| | | | 0.028 | 1309 | 20 | 56 | 24 |
| Frontal Pole | R | 16 | 0.043 | 1118 | 38 | 50 | -18 |
| Frontal Pole | L | 9 | 0.048 | 1078 | -18 | 48 | 4 |
| Temporal Pole | L | 76 | 0.031 | 1261 | -54 | 2 | -38 |
| Brainstem | L+R | 4 | 0.049 | 1067 | 12 | -16 | -16 |
| | | 38 | 0.044 | 1108 | -6 | -8 | -12 |
| | | | 0.047 | 1082 | 2 | -6 | -12 |
| | | 2 | 0.049 | 1067 | 6 | -8 | -10 |
| **Dystonic tremor patients only (N=23)** | | | | | | | |
| Cerebellum | L + R | 12594 | 0.001 | 2713 | -4 | -54 | -16 |
| | | | 0.001 | 2685 | 8 | -62 | -26 |
| | | | 0.001 | 2640 | 4 | -58 | -18 |
| | | 20 | 0.046 | 1099 | -16 | -76 | -56 |
| Frontal Pole | R | 554 | 0.032 | 1252 | 18 | 58 | 24 |
| | | | 0.032 | 1247 | 14 | 60 | 32 |
| | | | 0.034 | 1222 | 16 | 52 | 38 |
| Pallidum | L | 195 | 0.037 | 1188 | -16 | -8 | -8 |
| | | | 0.037 | 1187 | -6 | -10 | -12 |
| | | | 0.039 | 1163 | -14 | -4 | 4 |
| Thalamus L | | 39 | 0.046 | 1102 | -20 | -16 | 10 |
